# Anti–GM-CSF autoantibodies promote a “pre-diseased” state in Crohn’s Disease

**DOI:** 10.1101/2021.08.23.21262143

**Authors:** Arthur Mortha, Romain Remark, Diane Marie Del Valle, Ling-Shiang Chuang, Zhi Chai, Inês Alves, Catarina Azevedo, Joana Gaifem, Jerome Martin, Kevin Tuballes, Vanessa Barcessat, Siu Ling Tai, Hsin-Hui Huang, Ilaria Laface, Yeray Arteaga Jerez, Gilles Boschetti, Nicole Villaverde, Mona D. Wang, Ujunwa M. Korie, Joseph Murray, Rok-Seon Choung, Takahiro Sato, Renee M. Laird, Scot Plevy, Adeeb Rahman, Joana Torres, Chad Porter, Mark S. Riddle, Ephraim Kenigsberg, Salomé S. Pinho, Judy H. Cho, Miriam Merad, Jean-Frederic Colombel, Sacha Gnjatic

**Affiliations:** Precision Immunology Institute, Icahn School of Medicine at Mount Sinai, New York, NY, USA; Tisch Cancer Institute, Division of Hematology/Oncology, Department of Medicine, Icahn School of Medicine at Mount Sinai, New York, NY, USA; Human Immune Monitoring Center at Mount Sinai, New York, NY, USA; Department of Immunology, University of Toronto, Toronto, CA; Innate Pharma, Marseille, FR; Charles Bronfman Institute for Personalized Medicine, Department of Genetics, Icahn School of Medicine at Mount Sinai, New York, NY, USA; Hépato-Gastroentérologue, Hospices Civils de Lyon, Université Claude Bernard Lyon, FR; Department of Gastroenterology, Icahn School of Medicine at Mount Sinai, New York, NY, USA; Gastroenterology Division, Hospital Beatriz Ângelo, Loures, PT; Janssen R&D, Spring House, PA, USA; Naval Medical Research Center, Silver Spring, MD, USA; Division of Gastroenterology and Hepatology, Mayo Clinic, Rochester, MN, USA; Université de Nantes, Inserm, CHU Nantes, Centre de Recherche en Transplantation et Immunologie, UMR 1064, ITUN, 44000 Nantes, France; CHU Nantes, Laboratoire d’Immunologie, CIMNA, 44000 Nantes, France; i3S – Institute for Research and Innovation in Health, University of Porto, 4200-135 Porto, Portugal; Faculty of Medicine, University of Porto, 4200-319 Porto, Portugal; Institute of Biomedical Sciences Abel Salazar (ICBAS), University of Porto, 4050-313 Porto, Portugal

**Keywords:** Crohn’s Disease, innate lymphoid cells, macrophages, GM-CSF, autoantibodies

## Abstract

**Background & Aims:** Anti–GM-CSF autoantibodies (aGMAb) are detected in ileal Crohn’s Disease (CD) patients. Their induction and mode of action impacting homeostasis during, or prior to disease are not well understood. We aimed to investigate the underlying mechanisms leading to the induction of aGMAb, from functional orientation to recognized epitopes, for their impact on intestinal immune homeostasis and use as predictive biomarker for complicated CD.

**Methods:** Using longitudinally collected sera from active component US personnel, we characterize naturally occurring aGMAb in a subset of CD patients years before disease onset. We employed biochemical, cellular, and transcriptional analysis to uncover a mechanism that governs the impaired immune balance in CD years prior to diagnosis.

**Results:** Neutralizing aGMAb are specific to posttranslational glycosylations on GM-CSF, detectable years prior to diagnosis, and associated with complicated CD at presentation. Glycosylation and production of GM-CSF change in CD patients, altering myeloid homeostasis and destabilizing group 3 innate lymphoid cells. Perturbations in immune homeostasis precede the inflammation and are detectable in the non-inflamed CD mucosa of patients presenting with anti-GM-CSF autoantibodies.

**Conclusions:** Anti-GM-CSF autoantibodies predict the diagnosis of complicated CD, have unique epitopes, and impair myeloid cell homeostasis across the ILC3-GM-CSF-myeloid cell axis, altering intestinal immune homeostasis long before the diagnosis of disease.

## Introduction

Inflammatory bowel disease (IBD), sub-classified into Crohn’s disease (CD) and ulcerative colitis (UC), is an increasingly diagnosed chronic inflammatory pathology of the gastrointestinal tract, affecting 0.3% of the world’s population^1^. While both CD and UC affect the gastrointestinal tract, their causes remains puzzling and are likely multifactorial^2^. Genome-wide association studies (GWAS) provide strong support that IBD is a pathology driven by mono and multigenetic variations, but non-genetic and environmental factors are also considered as contributors to the heterogeneity of this disease^3, 4^. Identifying factors contributing to IBD development that may predict disease onset is therefore of high clinical relevance.

A key immunologic characteristic of IBD is the break in intestinal homeostasis, commonly manifested through insufficient barrier integrity, decreased immunologic tolerance, or excessive inflammation. The cytokine Granulocyte Macrophage-Colony Stimulating Factor (GM-CSF) reportedly plays a dual role in intestinal inflammation and was shown to have both protective and inflammatory properties in CD^5–9^. Group 3 innate lymphoid cells (ILC3) produce GM-CSF in the gut, which acts on myeloid immune cells to sustain immune homeostasis^10^. Derived from ILC3, GM-CSF promotes anti-bacterial and immunomodulatory functions in myeloid cells, to sustain the transcriptional stability of ILC3 via the metabolite retinoic acid (RA), further contributing to the generation and maintenance of immunosuppressive regulatory T cells (Treg)^10, 11^. However, in CD patients, myeloid cells induce differentiation of ILC3 into inflammatory group 1 ILC (ILC1) via Interleukin (IL)-12 and IL-23^12–14^. ILC3 or GM-CSF deficiency impairs anti-microbial immunity and mucosal homeostasis aligning with the identification of mutations in GM-CSF signaling in CD patients^6, 10, 15^. Administration of yeast-produced GM-CSF (sargramostim) improved CD and ameliorated symptoms^5, 16^. However, larger trials with sargramostim in CD failed to reach statistical significance, possibly due to high placebo group responses and suboptimal study design^17–19^.

Besides genetic defects in GM-CSF signaling, aGMAb may also contribute to CD in a subset of patients, and associate with ileal involvement, disease severity, higher relapse rates and complications^20–24^. Such autoantibodies can cause pulmonary alveolar proteinosis (PAP), resulting in alveolar macrophage-deficiency and increased lung pathologies^25^. Why PAP patients do not display intestinal pathologies and why CD patients with aGMAb do not show PAP-associated pulmonary symptoms remains unknown. Moreover, both the overproduction and absence of GM-CSF significantly increase the susceptibility to develop IBD, emphasizing the heterogeneity in CD pathogenesis, prompting us to investigate the biology of aGMAb prior to diagnosis in CD ^5–9, 26^.

Here, we analyzed >1800 sera, collected longitudinally from active component US military personnel over 10-years including subjects who eventually developed CD (n=220), UC (n=200), and personnel without CD or UC (HD, n=220) ^27^, along with controls with established CD or PAP. We analyzed titers, isotype profiles, epitopes, and functional consequences of aGMAb, using ELISA, neutralization, and reporter assays, and their effect on immune subsets in non-inflamed ileal CD biopsies by RNA sequencing, to establish mechanisms of pathology prior to disease and at its onset that could be exploited for personalized CD therapies.

## Methods

### Human specimen

Non-involved intestinal resection and involved intestinal resection samples were obtained from patients with CD undergoing ileal resection surgery at the Mount Sinai Medical Center (New York, NY) after obtaining informed consent. All protocols were reviewed and approved by the Institutional Review Board (IRB) at the Icahn School of Medicine at Mount Sinai (IRB 08-1236 and IRB HSM 13-00998). Stored pre-diagnosis serum samples were obtained from the Department of Defense Serum Repository, Silver Spring, MD, USA. 220 CD, 200 UC, and 200 healthy controls (HC) samples were provided, sampled at two to three time points prior to diagnosis of disease and one time point post diagnosis of disease^27^. Blood samples from six adult CD patients were collected at Porto University Centre Hospital, Portugal (CHUP). PAP control sera were kindly provided by Dr. Lanzavecchia (Institute for Research in Biomedicine, Switzerland). All patients were diagnosed with active CD and were naïve for treatment with biologics at the time of blood collection. Blood samples from healthy donors were used as controls. All participants gave informed consent about all clinical procedures and research protocols were approved by the ethical committee of both hospitals. Serum was collected from peripheral blood by centrifugation at 1620 x g for 10 minutes and stored at −80°C until analysis.

*Sample collection*: Resection samples and biopsies were cleared by a pathologist and placed in complete RPMI media on ice prior to processing. Patients diagnosed with UC or malignancies were excluded from the analysis.

*Isolation of PBMC*: Buffy coats obtained from the New York City Blood Center and PBMCs collected via a Ficoll gradient. PBMCs were washed, counted and resuspended in complete RPMI followed by consecutive experiments or further used for the enrichment of monocytes using magnetic anti-CD14-beads (purity of monocytes > 98%).

*Isolation of lamina propria leukocytes:* freshly removed ileal resections were washed in ice-cold PBS and scraped off of mucus. Epithelial cells were removed in HBSS free of calcium and magnesium, supplemented with EDTA (5mM) and HEPES (10mM) for 20-30 minutes at 37°C at 100rpm. Fibrotic tissue was removed and non-fibrotic mucosa minced and digested using Collagenase IV and DNase I (both Sigma) in HBSS (Ca^2+^Mg^2+^) 2% FBS for 30 minutes at 37°C at 100rpm agitation. Cell suspensions were filtered and live cells enriched using Percoll (80%:40%). The interphase was harvested, washed in FACS buffer (PBS, 5mM EDTA and 2% FBS) for subsequent use.

### Flow Cytometry and Mass Cytometry

*Aldefluor staining:* freshly isolated PBMCs or lamina propria leukocytes were washed twice in PBS and stained following the manufacturer protocol. Following Aldefluor staining, cells were washed in PBS and processed for surface staining.

*Surface staining:* Fc-receptors were blocked using Fc-block reagent (BD), following a 20 min surface staining with directly conjugated monoclonal antibodies. The following antibodies were used and purchased from BD, R+D, Biolegend and Miltenyi: anti-human CD45, anti-human HLA-DR, anti-human CD11c, anti-human CD14, anti-human CD1c, anti-human CD141, anti-human CD127, anti-human CD117, anti-NKp44, anti-CD161, anti-CD3, anti-CD19, anti-RORγt, anti-CD69. Following surface staining, cells were fix using the FOXP3 staining kit to stain for RORγt.

*Intracellular cytokine staining:* for intracellular cytokine staining of GM-CSF and IFN-γ, cells were re-suspended in complete RPMI supplemented with 10% FBS, 1% non-essential amino acids, 1% sodium-pyruvate, 1% L-glutamine, 1% penicillin-streptomycin and 1% HEPES. Media was further supplemented with Brefeldin A (for GM-CSF) or Brefeldin A, PMA (Sigma) and Ionomycin (Sigma) (for IFN-γ). Cells were incubated for 4h at 37°C and 5% CO2 prior to surface staining. Post surface staining, cells were fixed for 30minutes in Cytofix/CytoPerm (BD), washed in PBS containing 2% FBS, 5mM EDTA (FACS buffer) and 0.5% Saponin (Sigma). Anti-cytokine staining was performed for 30minutes at 4°C in the dark. Cells were washed and samples were analyzed on a BD LSR Fortessa II.

*Phospho STAT5 staining*: PBMCs or isolated LPL were resuspended in PBS and incubated with GM-CSF (10ng/ml) STAT5 phosphorylation was assessed using the BD PhosFlow Protocol for human PBMCs using PermBuffer II. Intracellular staining of pSTAT5 was performed using anti-human pSTAT5-Alexa647.

*Mass Cytometry*: Cells were stimulated at 1-3×10^6^/cells per reaction with recombinant GM-CSF (10ng/ml) for 20-30mins. Following stimulation cells were fixed for 30mins following the PhosFlow Protocol for human PBMCs using PermBuffer II (BD). Post fixation, groups of cells were barcoded using Cell-ID™ 20-Plex Pd Barcoding Kit and combinations of different metal isotope conjugated anti-CD45 antibodies. Pooled samples were stained with markers listed in SUPPLEMENTARY FIGURE 2A and analyzed on a Helios CyTOF system. Dimensionality reduction of CyTOF data was performed on the Cytobank software using the viSNE algorithm.

### Anti-GM-CSF ELISA, serum GM-CSF ELISA and Lectin ELISA

For anti–GM-CSF ELISAs, plates were coated with recombinant human GM-CSF (sargramostim), washed and blocked with TBST/BSA. Wells were incubated with 10-50µl of serum diluted in TBST followed by three washing steps. Anti–GM-CSF antibodies were detected by pan anti-human IgG HRP or isotype specific secondary antibodies. Substrate reaction was assessed using a plate reader at 550nm. Detection of lectin-binding motifs in the serum GM-CSF from CD patients and HD was performed by a lectin ELISA adapted from^76^. Prior to serum capture, serum from CD patients and HD was concentrated using Amicon® Ultra-2 mL Centrifugal Filters, to reach a final concentration of 4X. Microtiter plates (Maxisorp, Nunc) were coated with a mouse anti-human GM-CSF (DuoSet R&D Systems) in PBS buffer, overnight at room temperature (RT). Blocking was performed with Carbo-free blocking solution (Vector Labs) for 1h at RT. Plates were washed 5 times with PBS + 0.05% Tween 20 (PBST) before the CD/HD plasma samples in PBS containing 1% Carbo-free blocking solution (diluent solution) were added and incubated shaking at 200 rpm for 2h at RT. After washing the wells as described above, biotinylated mouse anti-human GM-CSF (DuoSet R&D Systems) was added and incubated for 2h shaking at 200 rpm at RT. Bound GM-CSF was detected using an HRP-conjugated streptavidin (DuoSet R&D Systems) incubated for 20 minutes and Tetramethylbenzidine substrate (DuoSet R&D Systems) was incubated for 20 minutes protected from dark. Reaction was stopped using H_2_SO_4_ and the amount of bound lectin was measured at 450 nm using a μQuant Microplate Reader (BioTek, Agilent).

*Antibody-binding assay*: GM-CSF was heat-denaturated in SDS containing buffer and used for anti–GM-CSF ELISAs. Binding-strength of anti–GM-CSF antibodies in patient sera was assessed using wash step with NaCl salt solutions of increasing concentrations (1-4M NaCl). Binding capacity was calculated as % of maximal binding.

*Stripping of glycosylation on GM-CSF*: GM-CSF was stripped using N-Glycosidase F, α2-3,6,8,9-Neuraminidase, Endo-α-N-acetylgalactosaminidase, β1,4-galactosidase and β-N-Acetylglucosaminidase (SIGMA) and according to the manufacturer’s recommendations.

*Native PAGE, SDS-PAGE/Western Blot, Lectin Blot*: Recombinant and stripped GM-CSF (sargramostim) (8µg/well) were separated on 15% resolving native polyacrylamide gels. Gels were stained with Coomassie G Brilliant Blue to confirm stripping. Proteins were then transferred to nitrocellulose membranes and membranes were blocked with 5% Non-Fat Dry Milk in Tris-Buffer Saline 0.1% Tween-20 (TBST) at 4°C. Membranes were then incubated with serum samples (diluted 1:100 in blocking buffer). Bound anti–GM-CSF antibodies were detected using anti-Human IgG AP at 1:1000 in TBST. Images of western blots have been cropped in Figure 1C and supplementary Figure 2B and 2C.

**FIGURE 1.**
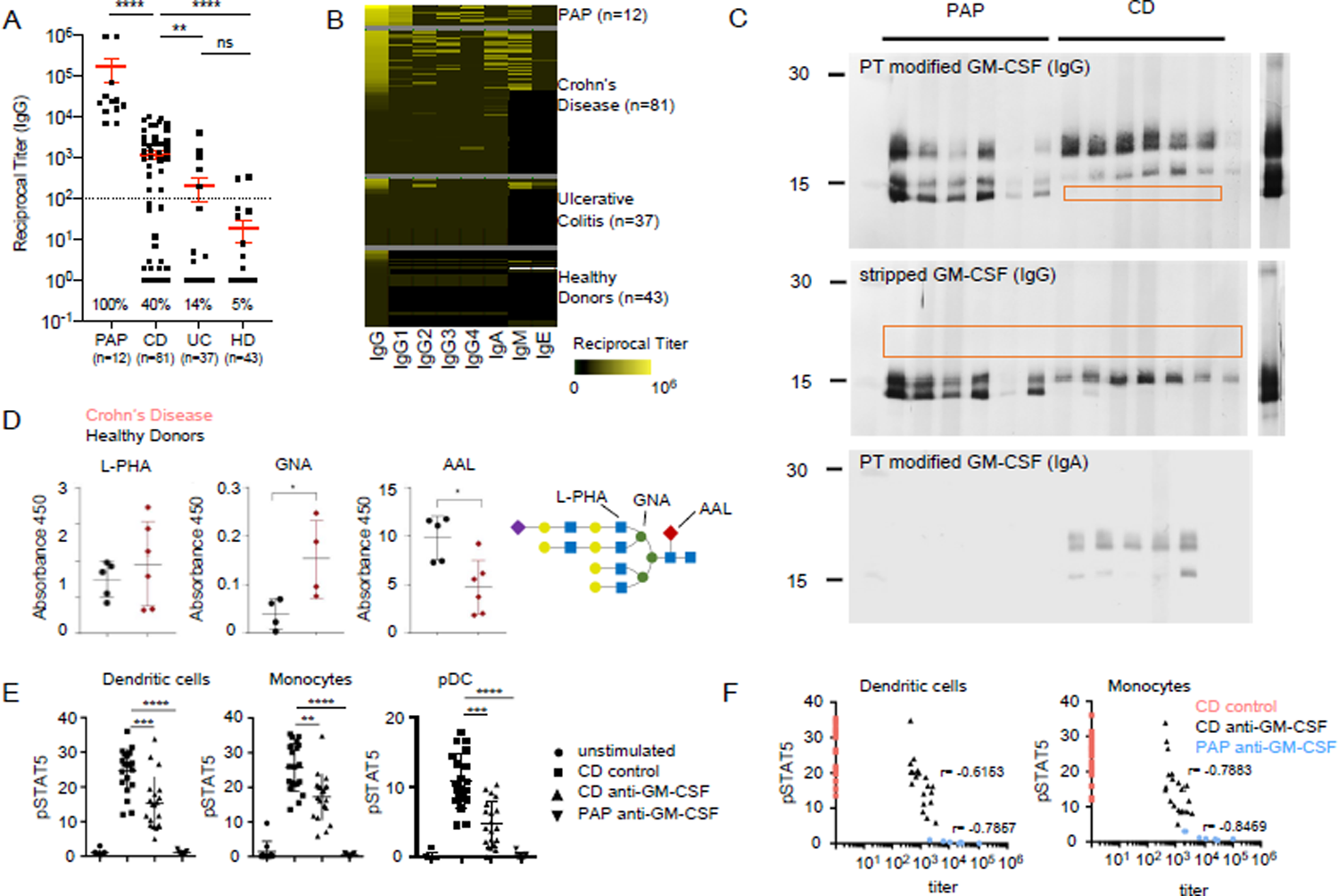
Characterization of aGMAb in CD patients. **A)** Reciprocal titers for total serum IgG aGMAb in sera of HD, PAP, CD and UC patients. **B)** Isotype profiles of aGMAb in PAP and CD patients. Horizontal rows represent patients and vertical rows indicate isotypes. **C)** Western blots probed with polyclonal sera from CD and PAP patients shows binding of anti-GM-CSF IgG and IgA to glycosylated (PT-modified) and stripped GM-CSF **D)** Levels of GNA, AAL and L-PHA binding to GM-CSF from healthy donors (black) and Crohn’s Disease (red) patients, normalized for the total levels of GM-CSF in each sample. Schematic representation of *N*-glycan highlighting lectin recognition. **E)** Plots show quantification of pSTAT5 signal in DC, MP and pDC either unstimulated or stimulated with GM-CSF pre-incubated with serum form the indicated patient groups. **F)** Loss in pSTAT5 correlates with aGMAb titers. One-way analysis of variance (ANOVA) Bonferroni’s multiple comparison test was performed. Mann-Whitney test *p-value <0.05

**FIGURE 2.**
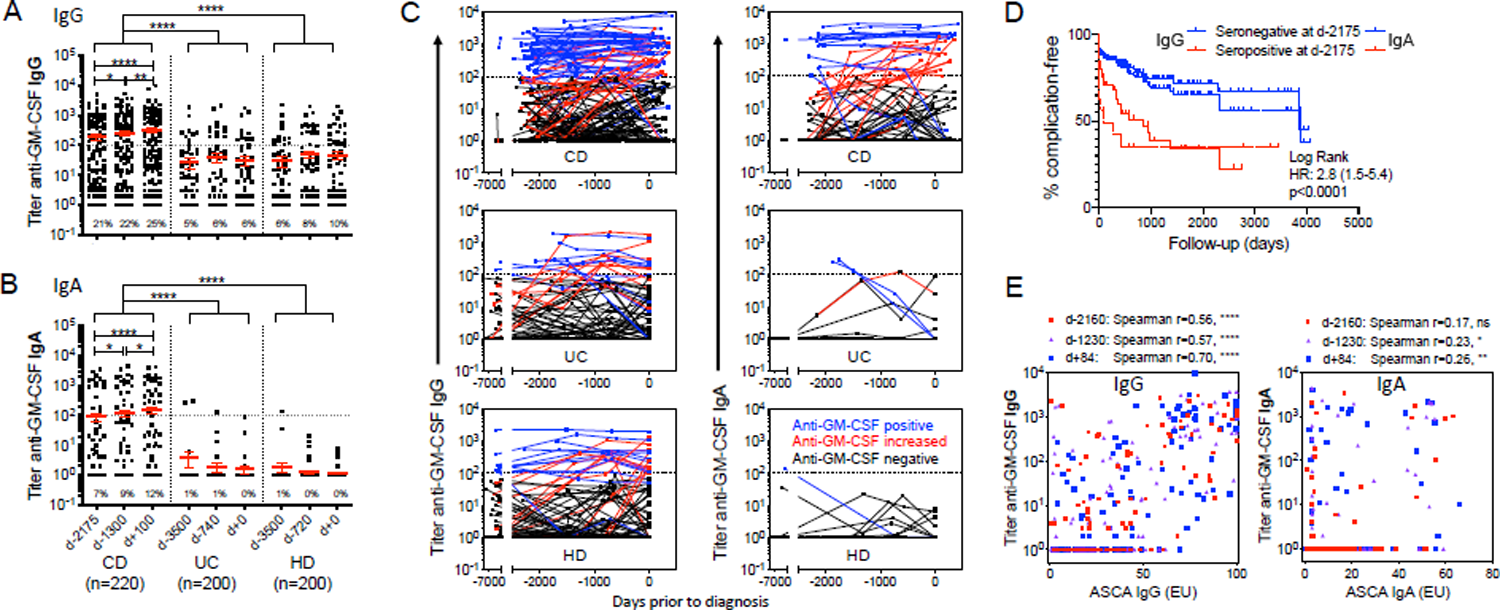
CD-specific aGMAb precede the onset of disease by years. **A)** and **B)** show reciprocal titers of aGMAb (IgG and IgA) in combined serum samples (training and validation cohort) at two time points prior to and one time point post diagnosis. **C)** Trajectory of aGMAb titers in CD patients. Blue lines indicate aGMAb^+^, black line aGMAb^-^ patients and red lines indicate sero-converters. **D)** Kaplan-Meier analysis with hazard ratio for developing complications after diagnosis in aGMAb^+^ (red) and aGMAb^-^ patients 6 years prior to disease. **E)** Correlations of aGMAb with ASCA IgG and ASCA IgA antibodies a different time points prior to diagnosis.

**FIGURE 3.**
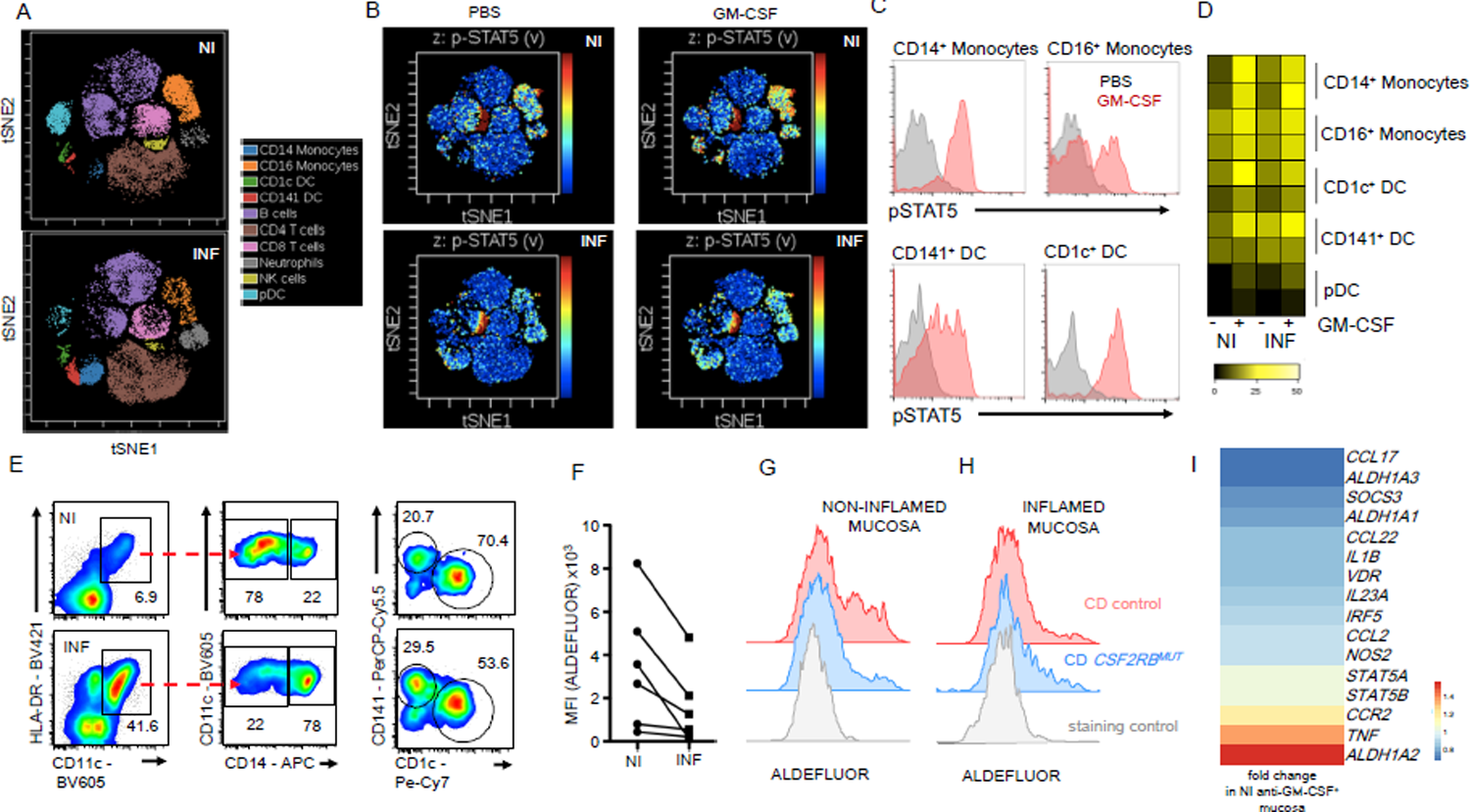
The inflamed CD mucosa shows impaired homeostatic functions in GM-CSF-responsive myeloid cells. LPL from NI and INF ileal mucosa were analyzed. **A)** ViSNE analysis shows distribution of leukocyte populations in NI and INF tissues indicating supervised annotation of populations. **B)** Stimulation of cells in **A)** with GM-CSF, followed by pSTAT5 measurement. Signal intensity is visualized in t-SNE plots (blue=low, red=high pSTAT5). **C)** Representative histograms show pSTAT5 levels in the indicated myeloid cell populations. **D)** Heat map shows pSTAT5 intensity across myeloid populations from **A)** and **B)**. **E)** DC subset and MP identification by floc cytometry. **F)** Mean fluorescence intensity (MFI) of ALDEFLOUR staining in CD45^+^CD11c^+^HLA-DR^+^ cells. ALDEFLUOR staining in CD45^+^CD11^+^HLA-DR^+^ cells in CD control and one *CSF2RB^MUT^* carrier from **G)** NI and **H)** INF tissue biopsies. **I)** Gene expression analysis of total tissue RNA from NI ileal tissues CD patients positive (n=14) of negative (n=27) for aGMAb. Heat map shows fold change in gene expression in aGMAb^+^ patients compared to aGMAb^-^ CD patients.

**FIGURE 4.**
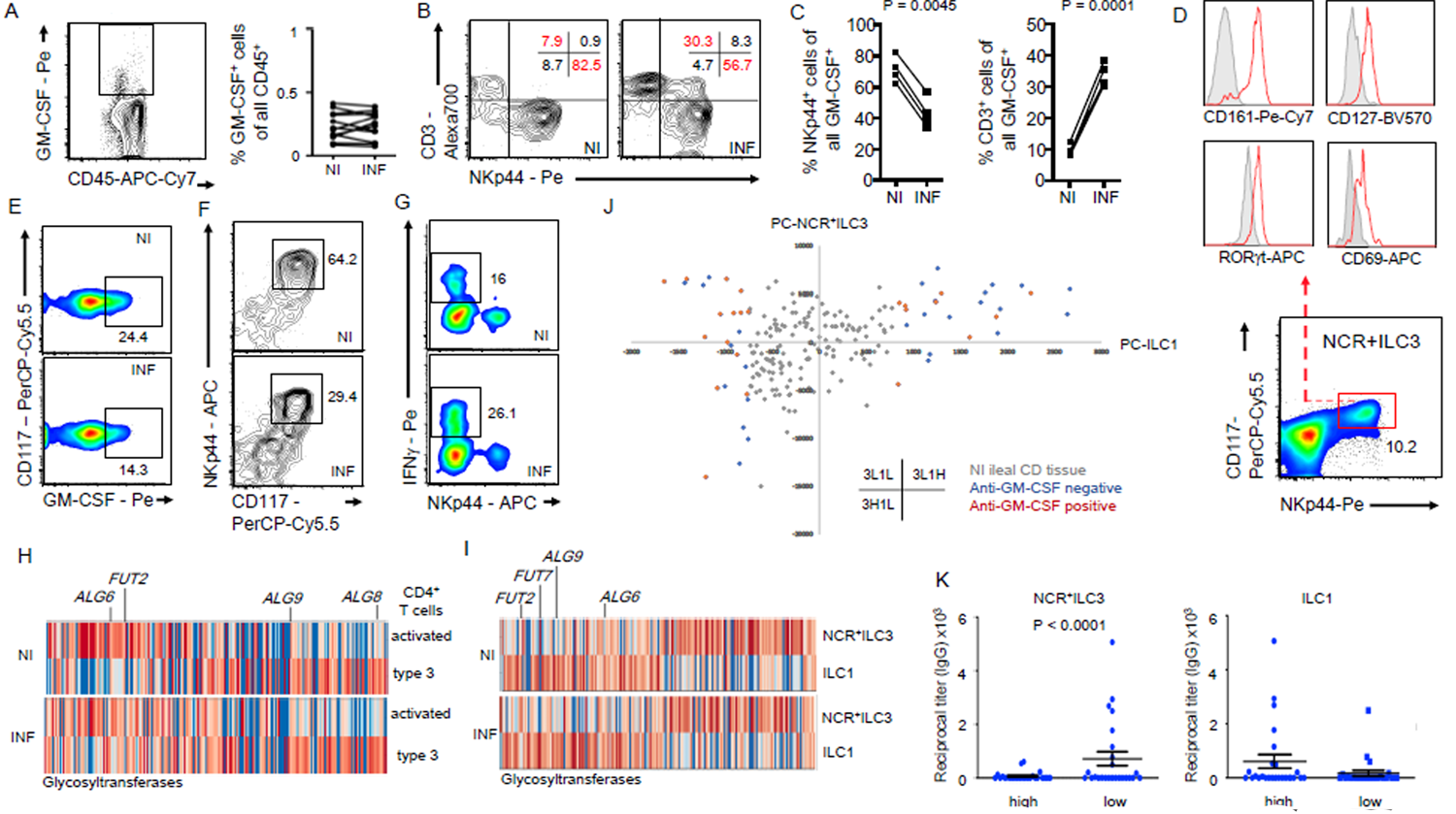
Innate and adaptive sources of intestinal GM-CSF in CD patients. **A)** shows GM-CSF^+^CD45^+^ cells the adjacent dot plot their frequency in NI and INF CD mucosa. **B)** GM-CSF^+^CD45^+^ cells were analyzed for CD3 and NKp44 expression in NI and INF CD mucosa. Numbers in gate represents percentages. **C)** Plots show percentages of NKp44^+^GM-CSF^+^ and CD3^+^GM-CSF^+^ cells. **D)** Expression of CD127, CD161, RORγt and CD69 on NKp44^+^ CD117^+^ cells. **E)** GM-CSF production by NCR^+^ILC3 (CD45^+^CD3^-^CD4^-^CD127^+^CD161^+^NKp44^+^CD117^+^), **F)** NCR^+^ILC3 frequencies and **G)** INFγ-producing ILC1 were quantified. Numbers in gates represent percentages. **H)** and **I)** show gene expression analysis of glycosyltransferase genes in the indicated population from INF and NI CD patients. **J)** Scatter plot shows PC1 values of ILC1-PC and NCR^+^ILC3-PC derived from PCA analysis of signature ILC1 and ILC3 genes against a sub-dataset (195 NI ileal CD and 121 NI ileal non-IBD samples) of the bulk mRNAseq samples from the MSCCR. Blue dots and red dots indicate anti-GM-CSF status. **K)** The top 25 samples with either the lowest and highest NCR^+^ILC3 or ILC1 signature gene expression were tested for aGMAb titers. Reciprocal titers are shown in plots.

*Lectin Blot and Western Blot:* The glycoprofile of recombinant forms of GM-CSF was evaluated by lectin blot. 2 μg of purified GM-CSF were separated as above and membranes blocked with BSA 4%. Phaseolus Vulgaris Leucoagglutinin (L-PHA), Maackia Amurensis Lectin II (MAL-II), Galanthus Nivalis Lectin (GNA) and Aleuria Aurantia Lectin (AAL) (each 2ug/mL) were used and bands visualized using the Vectorstain Elite ABC kit and ECL reagent. For western blot analysis 5μg of purified GM-CSF separated as above and detected using biotinylated GM-CSF antibody (0,1μg/mL R&D Systems). Target bands were visualized as above.

### Cloning, mutagenesis, expression and purification of GM-CSF

A double-stranded, C-terminal polyhistidine-tag (His-tag) containing *GM-CSF* DNA fragment was cloned into pIRESpuro2. Mutagenesis of putative glycosylation sites was performed using the Q5® Site-Directed Mutagenesis Kit and constructs of wild type and mutated *GM-CSF* stabely transfected into HEK293 cells (a kind gift by Dr. Goetz Erhardt). Cytokine secretion was validated using intracellular cytokine staining and ELISA as described above. Recombinant GM-CSF variants were purified using Ni-columns followed by western blot or assessment of bioactivity on U937 cells (a kind gift by Dr. Goetz Erhardt, University of Toronto).

### ILC gene signatures

ILC1: TBX21, IL12RB2, IL12RB1, IL2RB, GZMB, IFNG, FASLG, IL2RA, IRF8, GZMA, IKZF3, EOMES, ZNF683, GZMM ILC3: TNFSF13B, AHR, CCR6, CCL20, IL1R1, ID2, TOX2, NR1D1, IL22, LIF, RORC, KIT, IL23R, CSF2, IL7R, MAF

### Statistical Analyses

We used several visualization and clustering tools to describe the association of the investigated cells and analytes, e.g., scatter plot, heatmap, and t-distributed Stochastic Neighborhood Embedding (t-SNE). Continuous variables were summarized by disease groups (CD, UC, HD, and PAP) using standard descriptive statistics: median, interquartile range (IQR), mean, and standard deviation (SD) as appropriate. Antibody titers ≥ 100 were categorized as seropositive, and seronegative otherwise. The differences in analytes between the groups were assessed using Mann-Whitney, Student t-test, Kruskal-Wallis, and Analysis of variance (ANOVA) as appropriate. The differences in longitudinal antibody values between sampling time points were evaluated using paired t-test or Wilcoxon tests. We classified the trajectory groups based on the longitudinal antibody titers at earliest time point vs. follow-up time points: 1. Positive: titers ≥100 at baseline and follow-up time points; 2. Negative: titers <100 at baseline and follow-up time points; and 3. Increase: titers ≥100 and increasing at least four times compared to titers at previous time point(s). We explored the impact of prior antibody titers on development of severe CD using the Kaplan-Meier method with log-rank hazard ratio calculations. Data in the main figures show the combined data from a set of 320 patients (n=960 samples) used as a training cohort and another 300 patients (n=1200 samples) obtained and tested independently as a validation cohort (the data for training and validation cohorts is presented separately in supplemental figures). The model determined by the training cohort was then tested in the validation cohort, which performance metrics included sensitivity, specificity, and area under curve (AUC). The correlation between antibodies to GM-CSF and to ASCA or between GM-CSF autoantibody titers and pSTAT5 levels was assessed using Spearman’s correlation coefficient. The primary analyses were two-sided at the significant level of 0.05. We used the Bonferroni method to adjust for the situation of multiple testing.

## Results

### CD-associated aGMAb are distinct from those in PAP and UC

To assess the prevalence and characteristics of aGMAb in IBD, serum IgG titers against sargramostim, a yeast-produced recombinant GM-CSF, were tested in a cohort of patients with active CD (n=81) or UC (n=37), healthy donors (HD, n=43) and PAP patients (n=12). Among IBD patients, 40% of CD and 14% of UC patients displayed detectable levels of IgG aGMAb (titers >1/100) (FIG.1A). Titers detected in CD but not UC patients were significantly higher compared to HD, but lower when compared to PAP patients (FIG.1A). Only 3/53 sera from IBD patients with aGMAb reacted with other known autoantigens, emphasizing that the vast majority of aGMAb responses were antigen-specific (FIG.S1A). Sera showing broad non-specific antigen reactivity were excluded. Even though aGMAb titers differed between PAP and IBD patients, adjusted titers demonstrated comparable, relative avidity to GM-CSF (FIG.S1B).

We examined the isotypes and IgG subclasses of aGMAb in IBD vs. PAP patients. PAP-associated aGMAb were enriched in IgG1 and IgG4, isotypes virtually absent in IBD-associated aGMAb, were enriched in IgG2 and IgA. IgM and IgG3, but not IgE, were detectable in both PAP and IBD, with higher average IgM aGMAb in CD patients (FIG.1B). IBD-associated aGMAb were a specific marker for CD with ileal involvement and complicated disease, confirming previous studies (FIG.S1C-D and TAB.S1)^21–24^. Measuring aGMAb thus allows the discrimination of a subgroup of CD patients amongst all IBD patients.

Another major difference between IBD- and PAP-associated aGMAb was found in their epitopes. Peptide epitopes in PAP patients have previously been described in GM-CSF, but synthetic overlapping 20-mer linear peptides covering GM-CSF failed to react with CD-associated aGMAb (data not shown)^25^. Suspecting conformational epitopes, binding of aGMAb to denaturated GM-CSF was virtually absent using CD sera (FIG.S1E). Unlike PAP sera, CD sera did not react with GM-CSF in the absence of posttranslational modifications: only PAP sera bound bacterially produced recombinant GM-CSF, while CD sera required a eukaryotic GM-CSF product in order to react (data not shown). Analysis of yeast-produced GM-CSF by gel electrophoresis revealed three bands at ∼19.5 kDa, ∼16.5 kDa and ∼14.5 kDa (FIG.S1F). The highest band carried posttranslational glycosylations that were lost when enzymatically stripped (FIG.S1F). GM-CSF expressed in HEK293 cells lost this band pattern when all glycosylation sites were mutated to alanine (FIG.S1G). Posttranslational modifications of GM-CSF have previously been suggested as ideal antibody-recognition sites^28^. We assessed whether sera from CD patients would react with the different forms of yeast-derived GM-CSF. While PAP sera bound all bands of GM-CSF, CD-associated aGMAb bound larger glycosylated GM-CSF (FIG.1C, S1E-F). When sargramostim was enzymatically stripped of glycans, seroreactivity of CD to the 19.5 kDa band was lost but remained to the 16.5 kDa (FIG.1C), while PAP sera also recognized the 14.5 kDa band, which reflected the fully deglycosylated form of GM-CSF ( FIG.S1G)^25^. Similarly to IgG, IgA aGMAb from CD patients showed high specificity to glycosylation (FIG.1C). We confirmed specificity of CD-associated aGMAb using recombinant human GM-CSF produced in human cells, indicating that reactivity was not exclusive to yeast-specific modifications (data not shown). To evaluate commonalities in the glycan composition, yeast- and mammalian cell-derived GM-CSF, were resolved via SDS-PAGE and the glycoprofile of GM-CSF assessed via lectin blot. To determine commonalities in the glycosylation of GM-CSF, membranes were probed with L-PHA, MALII, GNA and AAL (FIG.S1H). Mammalian cell-derived GM-CSF displays a 25kDa glycoform reacting positive with L-PHA and AAL implicating complex branched and fucosylated N-glycans (FIG.S1I). The 16-17kDa band of mammalian cell-derived GM-CSF reacted with MALII and GNA, revealing high-mannose N-glycans and hybrid N-glycans with terminal sialylation (FIG.S1I). Yeast-derived GM-CSF reacted only with GNA suggesting high-mannose N-glycans as shared glyco-modification (FIG.S1I). These results demonstrate that mammalian-derived GM-CSF shares high mannose N-glycans with yeast-derived GM-CSF and suggest that CD-associated aGMAb selectively recognize these structures on correctly folded, native GM-CSF. To determine if glycosylations on GM-CSF differed between CD and HD, serum GM-CSF was captured and incubated with lectins. GNA binding was significantly higher on GM-CSF from CD patients, suggesting an increase in high mannose N-glycans on GM-CSF in CD (FIG.1D). While serum GM-CSF levels did not differ in CD and HD, decreased AAL binding indicated a decrease in the core fucose residues in CD (FIG.1D and FIG.S2A). The increase in mannose and decrease in the core-fucose structures suggest a CD-specific glycol-signature on GM-CSF.

### Neutralizing capacity of CD-associated aGMAb

The enriched abundance of binding of aGMAb to GM-CSF in CD patients (TAB.S1 and FIG.1A-B) supports the hypothesis that aGMAb possess neutralizing capacities^20^. We next determined whether aGMAb abrogate GM-CSFR signaling in monocytes, dendritic cells (DC) and plasmacytoid DC (pDC) of HC. Blood leukocytes stimulated with recombinant human GM-CSF *ex vivo* for 20min in the presence of PAP-sera (n=9), CD-sera negative for aGMAb (n=20), or CD-sera positive for aGMAb (n=20). Post stimulation, phosphorylation of STAT5 as measured by mass cytometry (TABLE.S2). Reduced STAT5 phosphorylation was observed in the presence of PAP-sera and CD-sera positive for aGMAb (FIG.1E and FIG.S2B-C). Stimulation of with GM-CSF did not result in pSTAT5 in T/B/NK cells across all groups (FIG.S2B). IL-3 stimulation on basophils was likewise unaffected by aGMAb^+^ CD-sera, suggesting unaltered common beta chain signaling (FIG.S2D). Decreased pSTAT5 correlated with aGMAb titers demonstrating neutralizing activity on circulating precursors of intestinal antigen-presenting cells (APC) (FIG.1F).

### CD-associated aGMAb precede the onset of complicated disease

The presence of neutralizing aGMAb in CD patients suggests that isotypes may contribute to disease pathophysiology and possibly etiology. In order to address this hypothesis, we tested sera obtained from the Department of Defense Serum Repository, prospectively collected from military service members during their annual routine medical examinations^29^. Some of these service members were eventually diagnosed with either UC or CD during the course of their service. Three to four longitudinal serum samples spanning up to 10 years, obtained prior, at and post diagnosis from 220 CD, 200 UC, and 200 matched individuals remaining healthy (HD), were analyzed in two independent runs (TAB.S3). Total IgG and IgA anti–GM-CSF ELISAs were performed for each time point of collection. Healthy military service members and service members diagnosed with UC had a similar 5-10% detection rate of anti-GM-CSF IgG, with low mean titers below the limit of significance (between 1/25 and 1/50), and nearly no anti–GM-CSF IgA detection (0-1%), without significant change by time point (FIG.2A-B and FIG.S3A-D). In contrast, IgG and IgA aGMAb were already found 6 years prior to CD diagnosis in 21% and 7% of samples, with additional patients seroconverting and mean titer increasing from 1/190 to 1/320 towards the time of diagnosis (FIG.2A-C and FIG.S3). At the time of diagnosis, IgA aGMAb were exclusively elevated in 12% of CD, while IgG were significantly more frequent (25%) compared to HD and UC (FIG.2A-B and FIG.S3C). Nearly all CD patients with detectable aGMAb 6 years prior to diagnosis maintained or increased their titers until the time of diagnosis (FIG.2C and FIG.S3E-F). Most (75%) anti–GM-CSF IgA co-occurred with IgG, while IgG to GM-CSF was more frequent and detected in 63% of cases in the absence of IgA.

Anti-GMAb in CD associated with ileal/ileocolonic involvement and complications within 100 days of diagnosis, with a 2.8 risk hazard ratio of having penetrating and/or stricturing disease or surgery at, or soon after clinical diagnosis (FIG.2D and FIG.S4A-D). Presence of IgA up to 6 years prior to diagnosis provided a predictor for CD development with >97% specificity and with sensitivity increasing from 15% to 21% as time to diagnosis decreased (AUC 0.6). The detection of aGMAb did not correlate with date of birth, sex, race, or year of sample acquisition, rendering this biomarker universal across patients (data not shown). Anti-*Saccharomyces cerevisiae* antibodies (ASCA) a common serological marker for IBD were shown to present prior to diagnosis using similar cohorts^3, 30, 31^. Anti-GMAb preceded the occurrence of anti-ASCA IgA and showed no significant correlations 2000 days prior to diagnosis (FIG.2E and TABLE.S4).

### The CD mucosa shows impaired homeostatic functions in GM-CSF-responsive myeloid cells

GM-CSF engages the heterodimeric GM-CSFR, composed of the GM-CSF binding alpha chain CD116 (*CSF2RA*) and the signal transducing common beta chain CD131 (*CSF2RB*) activating the transcription factor STAT5. To understand impairments in GM-CSFR signaling, we determined CD116 and CD131 expression across most hematopoietic cells in the inflamed (INF) and non-inflamed (NI) CD mucosa (TAB.S2B). While CD16^+^ or CD14^+^ monocytes, CD141^+^ DC, CD1c^+^ DC, pDC and neutrophils differed in their abundance, CD116 and CD131 expression and STAT5 phosphorylation remained unchanged between INF and NI CD tissues (FIG.3A-D and FIG.S5A-B). These findings suggest unperturbed GM-CSFR expression and responsiveness in the INF and NI CD mucosa.

GM-CSF stimulation controls essential myeloid functions including RA production. We next assessed the production of RA by APCs using ALDEFLUOR staining on HLA-DR^+^CD11c^+^ APCs from the INF and NI CD mucosa. APCs including CD14^+^ MP, CD141^+^ DC and CD1c^+^ DC showed decreased ALDEFLUOR staining specifically in the INF mucosa (FIG.3E-F, FIG.S5C-D). Monocytes and precursor DC continuously infiltrate the intestinal mucosa to differentiate into DC and MP and require GM-CSF for RA production^10, 15, 32, 33^. GM-CSF-stimulation of blood-derived CD14^+^ monocytes confirmed a GM-CSF-dependent increase in ALDEFLUOR staining comparable to staining in APCs from the NI CD mucosa (FIG.S5C-E). APCs isolated from a patient carrying a *CSF2RB^MUT^* allele revealed the requirement for GM-CSF in maintaining APC function even in NI tissues (FIG.3G-H). To determine if aGMAb alter GM-CSF-driven gene expression in the NI CD mucosa, expression of *STAT5A, STAT5B, IRF5, CCR2, CCL22, CCL17, VDR, SOCS3, ALDH1A1, ALDH1A2, ALDH1A3, TNF, NOS2, IL1B* was investigated in total tissue RNAseq data. Subdivision of patients based on their aGMAb status revealed decreased *ALDH1A1, ALDH1A3, CCL22, IL23A, VDR, CCL17, IRF5, CCL2, NOS2* and *IL1B* levels and increased *CCR2*, *TNF* and *ALDH1A2* expression in the presence of aGMAb (FIG.3I). These data suggest that GM-CSF regulates homeostatic gene expression incl. RA production in the NI CD mucosa, implicating an aGMAb-driven perturbation of homeostasis even prior to inflammation.

### T cells and ILC3 contribute to the pool of GM-CSF in the non-inflamed and inflamed CD mucosa

Assessment of spontaneously released GM-CSF by CD45^+^ cells was comparable between INF and NI CD tissues (FIG.4A), in line with unchanged serum GM-CSF levels in CD patients (FIG.S2A). A characterization of GM-CSF-producing cells in the NI CD ileal mucosa revealed that 80% were NKp44^+^CD3^-^ innate lymphoid cells (ILC), while the remaining 20% composed of CD3^+^ T cells, or cells lacking both markers (FIG.4B). Interestingly, the number of GM-CSF-producing T cells increased in the INF CD mucosa while the number of GM-CSF^+^NKp44^+^ cells decreased (FIG.4B-C). GM-CSF-secreting NKp44^+^ cells co-expressed CD117, CD127, CD161, CD69 and the transcription factor *Retinoic acid-related Orphan Receptor* (ROR) *gamma* (γ) t (FIG.4D), identifying them as natural cytotoxicity receptor (NCR)^+^ group 3 ILC (NCR^+^ILC3)^34^. Analyzing NCR^+^ILC3 numbers and cytokine release revealed lower levels of GM-CSF production and GM-CSF producers in the INF mucosa (FIG.4E-F and FIG.S5F). Despite of increased GM-CSF^+^ T cell frequencies, a lower per cell output was observed in these cells (FIG.4C and FIG.S5F). We next determined if NCR^+^ILC3 increasingly differentiated into group 1 ILCs (ILC1) as reported in CD^12, 14^. ILC1s were increased in the INF mucosa releasing higher levels of interferon(IFN)-γ (FIG.4G)^35, 36^. These findings demonstrate changes in the source and levels of GM-CSF in the INF CD mucosa, accompanied by decreased NCR^+^ILC3 and increased ILC1 counts (FIG.4E-G). The aberrantly glycosylated GM-CSF observed in CD sera (FIG.1D), inspired the analysis of glycogene-expression in GM-CSF^+^ leukocytes of the INF and NI CD mucosa. Published scRNA-Seq data from CD patients was analyzed for the expression of glycosyltransferases in T cells, NCR^+^ILC3s and ILC1^37^. Glycogene signatures differed in T cells and NCR^+^ILC3s between INF and NI mucosa, implicating the production of differential glycovariants of secreted proteins including GM-CSF. T cells and NCR^+^ILC3 in the INF mucosa displayed an upregulation of mannose-related (*ALG6*, *ALG8*, *ALG9*), fucosylation-related (*FUT2, FUT7*) and α2,3-sialylation-related glycogenes, when compared to the NI mucosa, in line with previous reports (FIG.4H-I)^38–42^. Noteworthy, substrate availability and modulation of glycosyltransferases expression may collectively, or individually result in aberrantly glycosylated proteins including GM-CSF. The decreased availability of NCR^+^ILC3-derived GM-CSF was reported to reduce myeloid RA production in mice, aligning with our findings in CD patients (FIG.3E-G and FIG.S5C-F)^10, 43^. Importantly, myeloid-derived RA prevents the accumulation of inflammatory ILC1, suggesting that aGMAb perturb the equilibrium of tissue-resident ILCs in pre-diagnostic CD patients through the modulation of GM-CSF-dependent myeloid RA production^29, 30^. To determine if the presence of aGMAb coincided with altered NCR^+^ILC3 and ILC1 gene signatures, bulk mRNA-Seq data from NI ileal CD biopsies (n = 191) and non-IBD controls (n = 121) was re-analyzed for ILC1 and ILC3 signature gene expression. Samples were displayed based on their PC-NCR^+^ILC3 and PC-ILC1 signatures gene expression profiles and revealed groups with high NCR^+^ILC3 and ILC1 (3H1H), low NCR^+^ILC3 and ILC1 (3L1L) and low NCR^+^ILC3 and high ILC1 signatures (3L1H) (FIG.4J). We next evaluated aGMAb titers across the most differential PC signatures and found aGMAb enriched in groups low in NCR^+^ILC3 signature gene expression (FIG.4J-K), suggesting an association of aGMAb with lower NCR^+^ILC3 genes signatures in NI CD tissues.

### Unmodified GM-CSF as a potential way to restore homeostatic functions of GM-CSF

Enzymatically stripped and genetically engineered GM-CSF, lacking posttranslational glycosylations remained biologically active and initiated phosphorylation of STAT5 (FIG.S6A-C). We hypothesize that GM-CSF devoid of glycosylations would render the cytokine capable of escaping neutralization by CD-associated aGMAb. Freshly isolated PBMCs stimulated with glycosylated and deglycosylated GM-CSF in the presence or absence of aGMAb were analyzed for STAT5 phosphorylation. The neutralizing effects of CD-associated aGMAb were recapitulated for glycosylated GM-CSF but reverted when enzymatically stripped GM-CSF was used (FIG.S6A-D).

## DISCUSSION

Here, we report the presence of aGMAb in the sera of CD patients years before diagnosis, and propose that these antibodies contribute to the pathophysiology of CD. We demonstrated IgG2- and IgA-skewed aGMAb isotypes in CD patients, suggesting an origin within the intestinal mucosa. Anti–GM-CSF autoantibodies were associated with ileal disease location, were present up to 6 years before diagnosis in asymptomatic subjects developing CD, and predicted complications at disease presentation (HR=2.9 by log rank, p<0.001). IgA aGMAb, were an exclusive hallmark of CD, and blocked GM-CSFR signaling by binding to GM-CSF glycovariants. This in turn impaired communication of ILC3 and myeloid cells in the NI CD mucosa at steady-state, resulting in significantly lower expression of an ILC3-specific gene signature in the presence of aGMAb. Furthermore, GM-CSF target gene expression in the NI mucosa of CD patients is reduced in the presence of aGMAb, identifying a subgroup of individuals at high risk of developing complicated CD through alterations of the intestinal immune balance. Importantly, GM-CSF in CD patients is aberrantly glycosylated via an altered gene expression of glycosyltransferases in GM-CSF-producing ILC3 and T cells, suggesting immunogenicity of post-translational epitopes recognized by aGMAb. Together, these results support a novel mechanism of disease pathophysiology that may be exploited for developing personalized CD-preventive and therapeutic strategies.

Myeloid cells of the INF CD mucosa displayed markedly reduced GM-CSF-dependent RA production, associated with reduced NCR^+^ILC3 numbers, lower GM-CSF output and altered glycogene expression. Abrogating GM-CSF-mediated signaling altered myeloid cell function and increased ILC1 signature gene expression even in NI tissues, suggesting a role for aGMAb in abrogating homeostasis^13, 14, 34, 36, 44^. Enrichment in lower ILC3 signatures in aGMAb^+^ CD patients suggest a GM-CSF-dependent myeloid regulation of the ILC3/ILC1 balance. Autoreactive aGMAb thus promote the accumulation of inflammatory ILC1 by disrupting the NCR^+^ILC3-myeloid cell circuit. We suggest that CD-associated aGMAb promote immune imbalance during the years-long “pre-diagnostic” period of CD by altering GM-CSF-dependent homeostasis, a hypothesis supported by our data that demonstrates reduced GM-CSF-dependent gene expression even in the NI CD mucosa of aGMAb^+^ patients.

There is rising interest in identifying the preclinical phase of CD where prevention strategies may eventually be pursued^45^. Measuring aGMAb adds to the list of predictive biomarkers which have been recently identified, such as microbial antibodies^46^ and intestinal permeability^47^. One specific interest is that aGMAb are associated with complicated CD disease at onset and thus may one day help identify relevant candidates for disease prevention at its earliest phase.

The differential glycosylation of GM-CSF and altered glycosyltransferase expression suggest that new (glyco)epitopes on GM-CSF may promote the development of aGMAb. Engineering of GM-CSF variants with functional activity but capable of circumventing aGMAb may provide a way to re-establish homeostasis, or delay disease progression. Our data calls for revisiting the use of GM-CSF in clinical trials with modified versions of active GM-CSF not prone to antibody neutralization, with careful pre-selection of patients based on their aGMAb profiles. Our findings demonstrate an intriguing mechanism for the development of complicated CD and allow the identification of these patients using a predictive serological biomarker. Collectively, these findings open new roads for the precise diagnosis, classification, and personalized treatment of CD patients.

### Grant support

A.M. is supported by a CIHR-Project Grant (388337), a NSERC-Discovery Grant (RGPIN-2019-04521). A.M. is the Tier 2 Canadian Research Chair in Mucosal Immunology and supported by the Tier 2 CRC-CIHR program. S.S.P. acknowledges Portuguese funds through the Portuguese Foundation for Science and Technology (FCT) in the framework of the project POCI-01-0145-FEDER-028772 and support by the Broad Medical Research Program at the Crohn’s and Colitis Foundation of America; the International Organization for the study of Inflammatory Bowel Disease, and the Portuguese Group of Study in IBD (GEDII) for funding. S.S.P. also acknowledges the US Department of Defense, US Army Medical Research Acquisition Activity, FY18 Peer Reviewed Medical Research Program Investigator-Initiated Research Award (award number W81XWH1920053). I.A. thanks FCT for funding [SFRH/BD/128874/2017]. S.L.T. acknowledges funding through the Dr. Edward KETCHUM Graduate Student Scholarships at the University of Toronto and the Canada Graduate Scholarships – Master’s (CGS-M) award. M.W. is support by a UROP fellowship by the Department of Immunology at the University of Toronto. J.H.C. acknowledges funding from U01 DK062429, U01 DK062422, R01 DK106593, and the Sanford Grossman Charitable Trust. S.G and J.F.C. are supported through U01 DK124165. S.G. is additionally supported by grants U24 CA224319. Funding and support of the PREDICTS study platform was provided through a Cooperative and Research Development Agreement with direct contributions by Janssen Pharmaceuticals, Prometheus Laboratories and the Naval Medical Research Center (NCRADA number NMR-11-3920). The mass cytometry instrumentation at the Human Immune Monitoring Center was obtained with support from S10 OD023547 and P01 CA196521.

## Disclosure

The University of Toronto and the Mount Sinai Hospital (NY) have collectively filed a patent application listing S.G., A.M., J.F.C., M.M. and R.R. as inventors that is related in part to this publication. S.G. reports consultancy and/or advisory roles for Merck, and OncoMed and research funding from Bristol-Myers Squibb, Genentech, Janssen R&D, Pfizer, Takeda, and Regeneron. JFC reports receiving research grants from AbbVie, Janssen Pharmaceuticals and Takeda; receiving payment for lectures from AbbVie, Amgen, Allergan, Inc. Ferring Pharmaceuticals, Shire, and Takeda; receiving consulting fees from AbbVie, Amgen, Arena Pharmaceuticals, Boehringer Ingelheim, BMS, Celgene Corporation, Eli Lilly, Ferring Pharmaceuticals, Galmed Research, Genentech, Glaxo Smith Kline, Janssen Pharmaceuticals, Kaleido Biosciences, Imedex, Immunic, Iterative Scopes, Merck, Microba, Novartis, PBM Capital, Pfizer, Sanofi, Takeda, TiGenix, Vifor; and hold stock options in Intestinal Biotech Development.

## Author Contributions

AM, RR, DMDV, LSC, ZC, IA, CA, JG, JM, KT, SLT, IL, AR, YAJ, GB, NV, MW performed experiments, sample processing data acquisition and analysis. JM, RSC, TS, RML, SP, CP, MSR, EK, SSP, JHC, MM, JFC, AM and SG provided guidance, facilitated sample acquisition or coordinated this study. AM and SG wrote the manuscript.

## Data transparency statement

The data that supports the findings of this study are available from the corresponding authors upon reasonable request.

---

**FIGURE S1.**
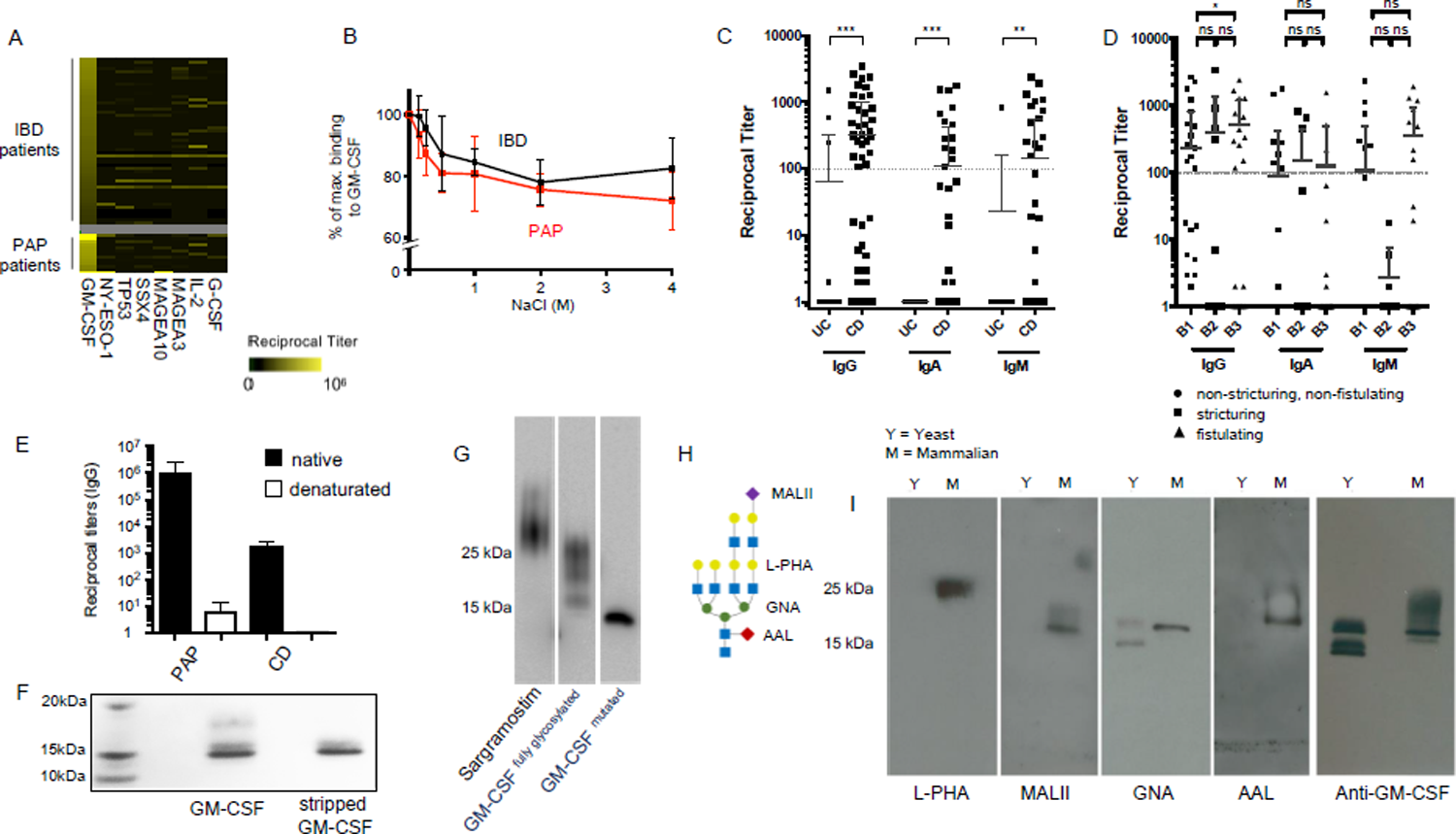
Specificity and affinity of anti–GM-CSF autoantibodies in CD patients. **A)** ELISA specific to cytokines (G-CSF, IL-2 and GM-CSF), nuclear antigens and unrelated autoantigens was performed using sera from PAP patients and IBD patients. Sera simultaneously reacting against GM-CSF and other antigens were excluded from the study. **B)** Binding avidity of aGMAb was determined using anti–GM-CSF ELISA with adjusted PAP and IBD sera. Post incubation of plates with anti–GM-CSF sera, wells were washed with buffers containing increasing amounts of NaCl. Signal detected by the ELISA plate reader post washing with high salt buffer was normalized to signals obtained in regular ELISAs. The binding strength of PAP sera (black) and IBD sera (red) are displayed. **C)** Anti–GM-CSF ELISA were performed using CD, UC and HC sera. Secondary antibodies recognizing IgG, IgA and IgM were used to identify enrichment of aGMAb in CD patients. **D)** Sera from CD patient were tested for their association with one of the three behavioral stages described in the Montreal Classification. **E)** Bar graph shows titers of aGMAb ELISA on native and denaturated GM-CSF using sera from PAP and CD patients. **F)** Native PAGE of GM-CSF (sargramostim) and stripped GM-CSF stained with Coomassie Brilliant Blue. **G)** Western blot of sargramostim, recombinantly expressed human GM-CSF purified from HEK293 cells stably secreting wild type human GM-CSF, or human GM-CSF mutated to lack glycosylations. **H)** Scheme adjacent to plot shows exemplified positioning of the evaluated N-linked glycosylations. **I)** Characterization of N-glycosylation of yeast (Y)- and mammalian (M)-produced GM-CSF by lectin blot for L-PHA, MALII, GNA and AAL. Western blot for GM-CSF confirms the position and molecular weight of the recombinant protein.

**FIGURE S2.**
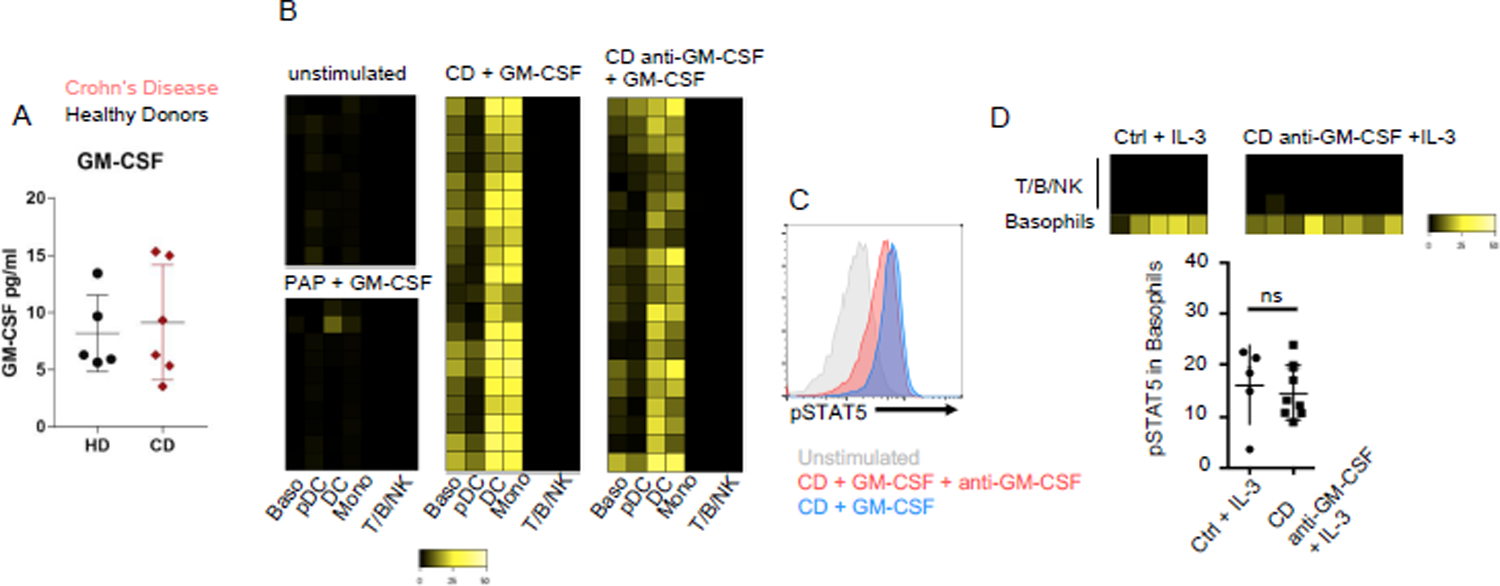
Neutralization of GM-CSF by aGMAb from CD patients. **A)** Total serum GM-CSF concentration in healthy donor (HD) or CD patients. **B)** PBMCs were isolated from a buffy coat. For every tested serum, 2 million PBMCs were seeded into the well of a 96 well plate. Cells were either left unstimulated or stimulated with rhGM-CSF for 20 minutes. GM-CSF-stimulated samples were pre-incubated with either serum from aGMAb^-^ CD patients, aGMAb^+^ CD patients or PAP patients. Cells were fixed after stimulation, barcoded prior to surface staining, then pooled and stained for surface marker and intracellular pSTAT5. Phosphorylation of STAT5 was analyzed in the indicated populations. Heat maps show signal intensity of anti-pSTAT5 staining in yellow color code for individual patients (lanes) within the indicated population (row). **C)** Representative pSTAT5 staining in monocytes either left untreated (grey), or stimulated with GM-CSF in the presence of control CD serum (blue) or CD serum containing aGMAb (red). **D)** PBMCs were stimulated in vitro with rhIL-3 in the presence or absence of the indicated sera for 20 minutes. Cells were washed, barcoded, pooled and then stained with surface antibodies follow by Intracellular staining with antibodies against phosphorylated STAT5. Mass cytometry was performed and intensity of pSTAT5 was visualized using heat maps. Scatter plots show quantification of IL-3 mediated STAT5 phosphorylation in Basophils in the presence aGMAb^-^ or aGMAb^+^ CD sera.

**FIGURE S3.**
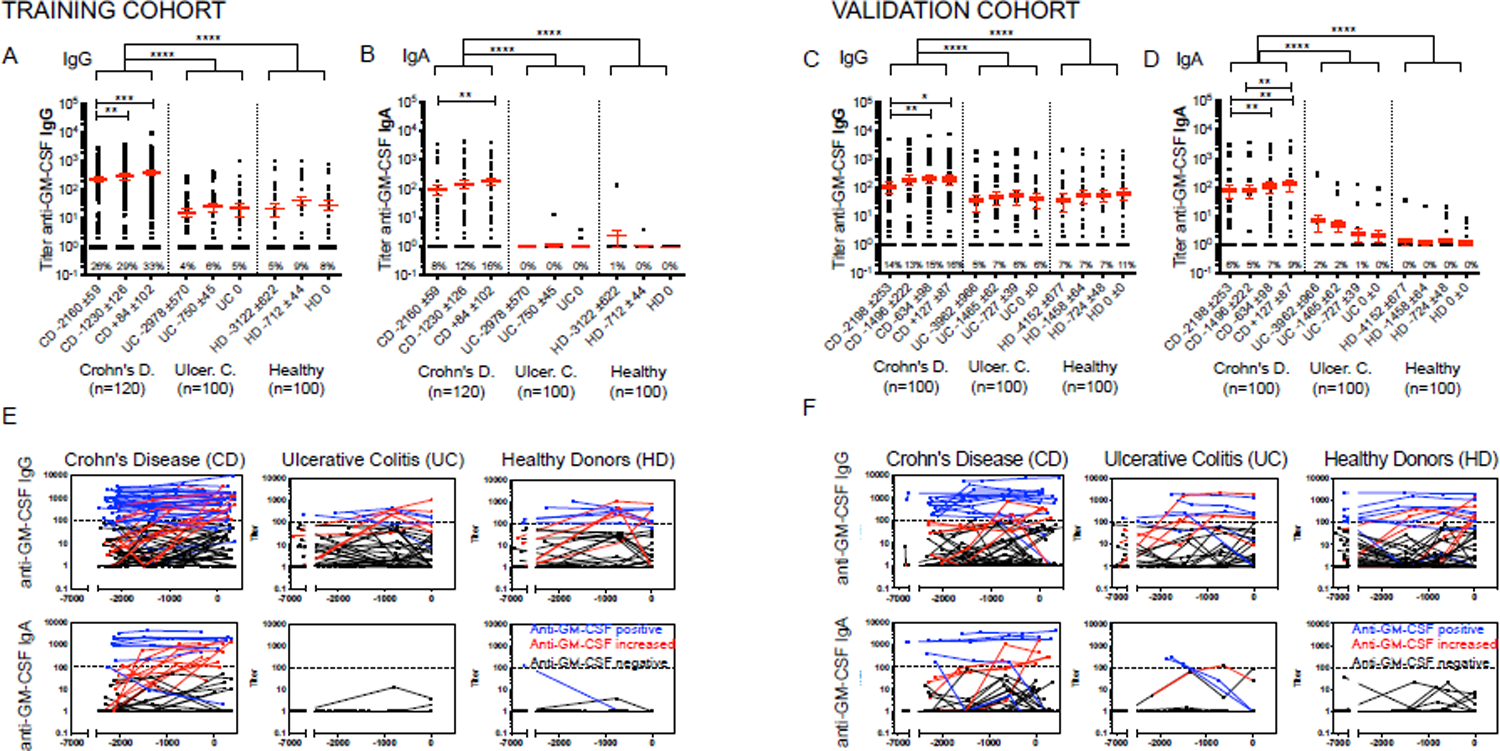
Anti–GM-CSF autoantibodies precede the onset of Crohn‘s Disease. **A-D)** Anti–GM-CSF ELISA was performed using serum samples obtained from CD patients, UC patients and HD. Sera were obtained at two (training cohort) and three (validation cohort) time points prior to diagnosis of disease and one time point after diagnosis of disease. Titers of anti–GM-CSF IgG and IgA were determined for each time point. Trajectory of aGMAb titers in CD, UC and HD across different time points are displayed in **E)** and **F)**. Blue line indicate patient tested positive at the earliest time point of collection. Red lines indicate sero-converter, while black lines indicate patients without aGMAb.

**FIGURE S4.**
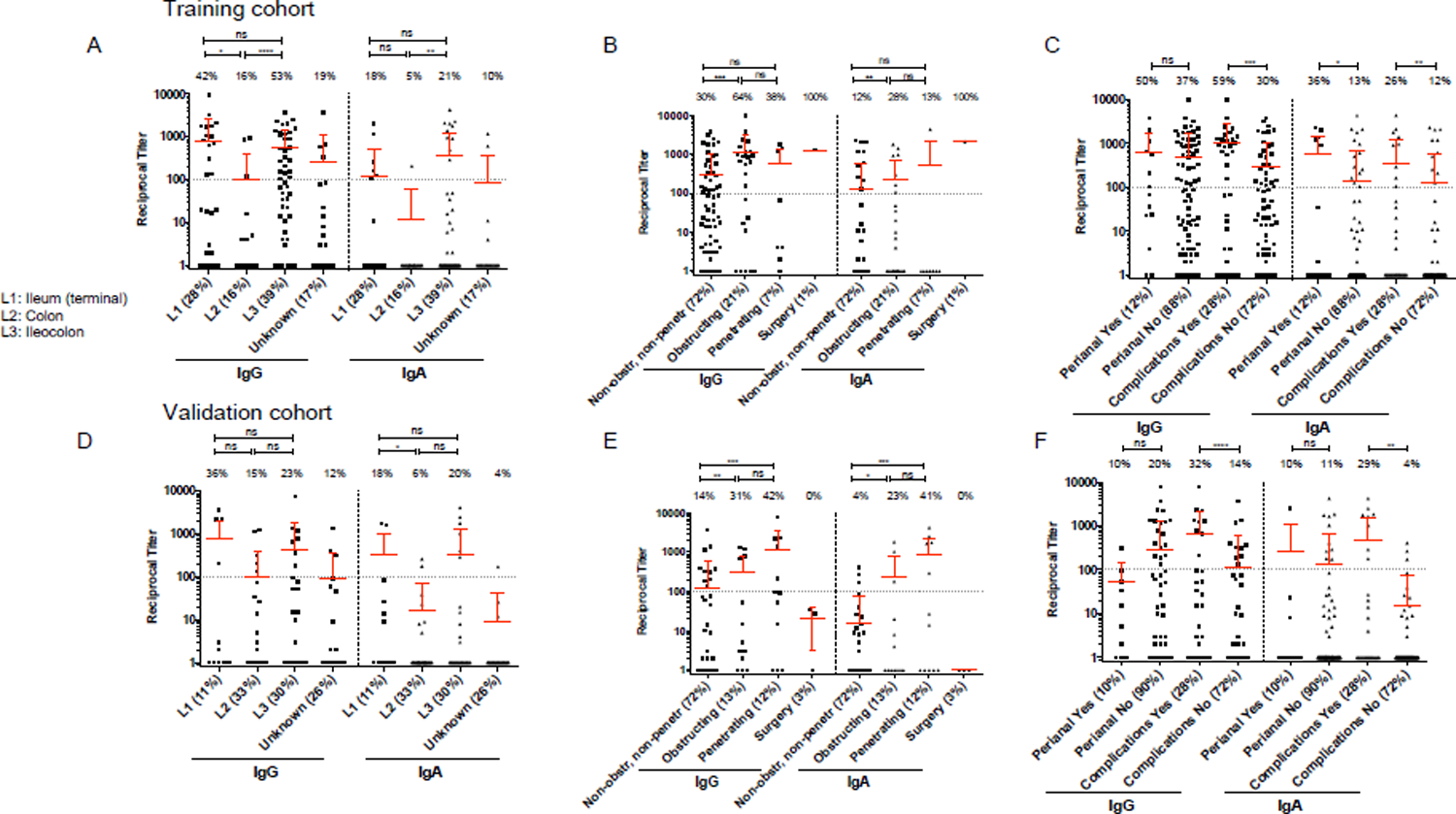
Anti–GM-CSF autoantibodies determine disease location and disease severity in two independent cohorts. **A-F)** Serum samples described in supplementary table 3 were analyzed for the association of IgG and IgA with disease location, obstruction, penetrance, surgery, perianal involvement and complications. A trainings cohort was established **A-C)** and compared to a validation cohort **D-F).**

**FIGURE S5.**
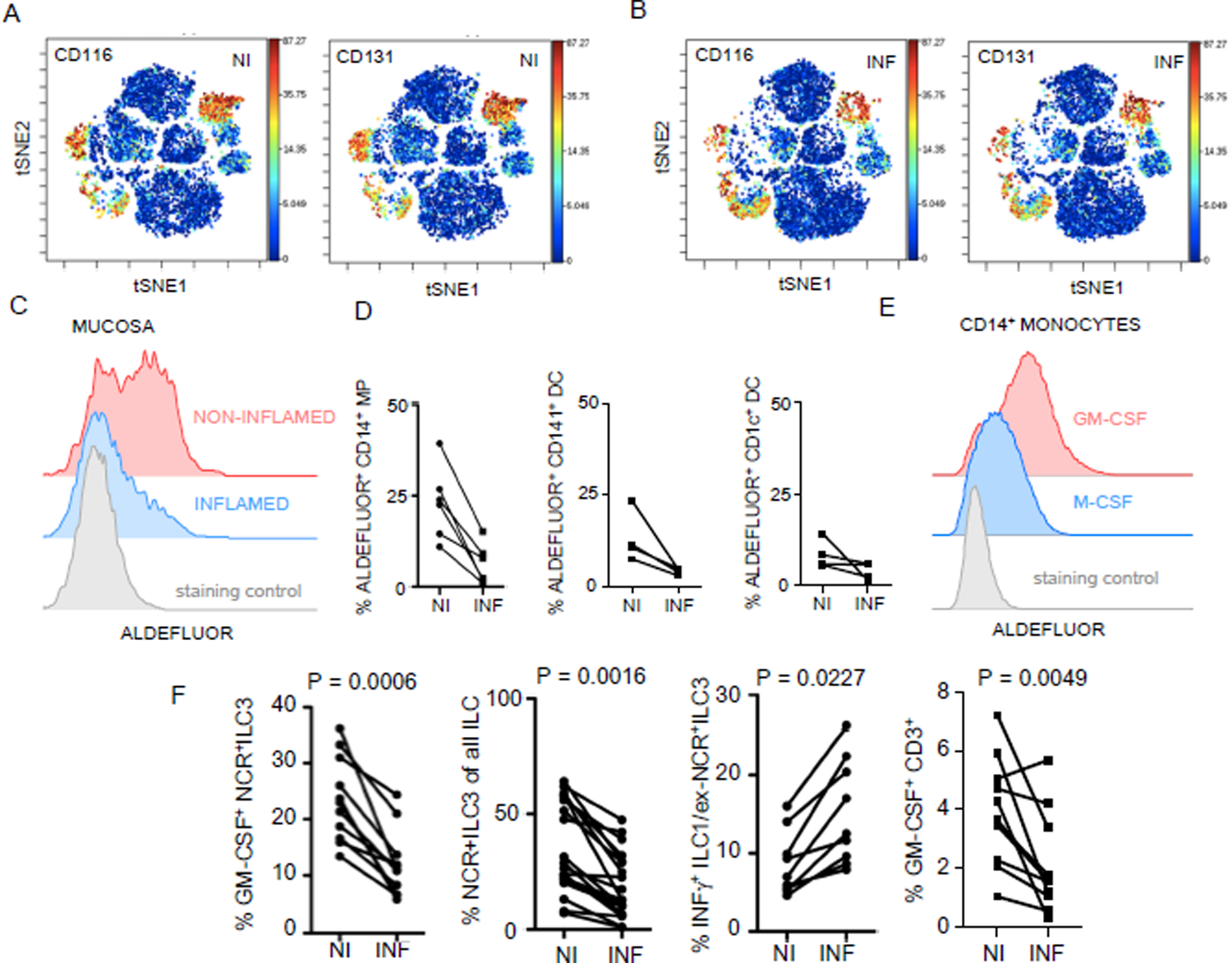
The inflamed CD mucosa shows intact GM-CSFR expression but reduced homeostatic, GM-CSF-dependent myeloid functions. **A)** and **B)** show CD116 and CD131 expression intensity across all leukocyte populations in NI **A)** and **B)** INF tissues identified by t-SNE analysis. Scale adjacent to plots indicates signal intensity. **C)** Histogram of representative ALDEFLOUR staining on intestinal macrophages from the NI (red) and INF (blue) CD mucosa. **D)** Plots show percentages of ALDEFLUOR staining^+^ MP, CD141^+^DC and CD1c^+^DC. **E)** Blood CD14^+^ monocytes were cultured in GM-CSF or M-CSF. Cells were analyzed for RA production using ALDEFLUOR staining 5 days later. **F)** Plots show quantification of GM-CSF^+^ NCR^+^ILC3 cells, relative abundance of NCR^+^ILC3s among all ILCs, IFNγ^+^CD45^+^CD3^-^CD4^-^CD127^+^CD161^+^NKp44^-^ ILC1/ex-NCR^+^ILC3 cells and GM-CSF^+^CD3^+^ cells. Datasets shown are representative of 9-20 individual ileal CD resections. Statistical analysis was performed using paired student’s t-test. P values are indicated adjacent to datasets.

**FIGURE S6.**
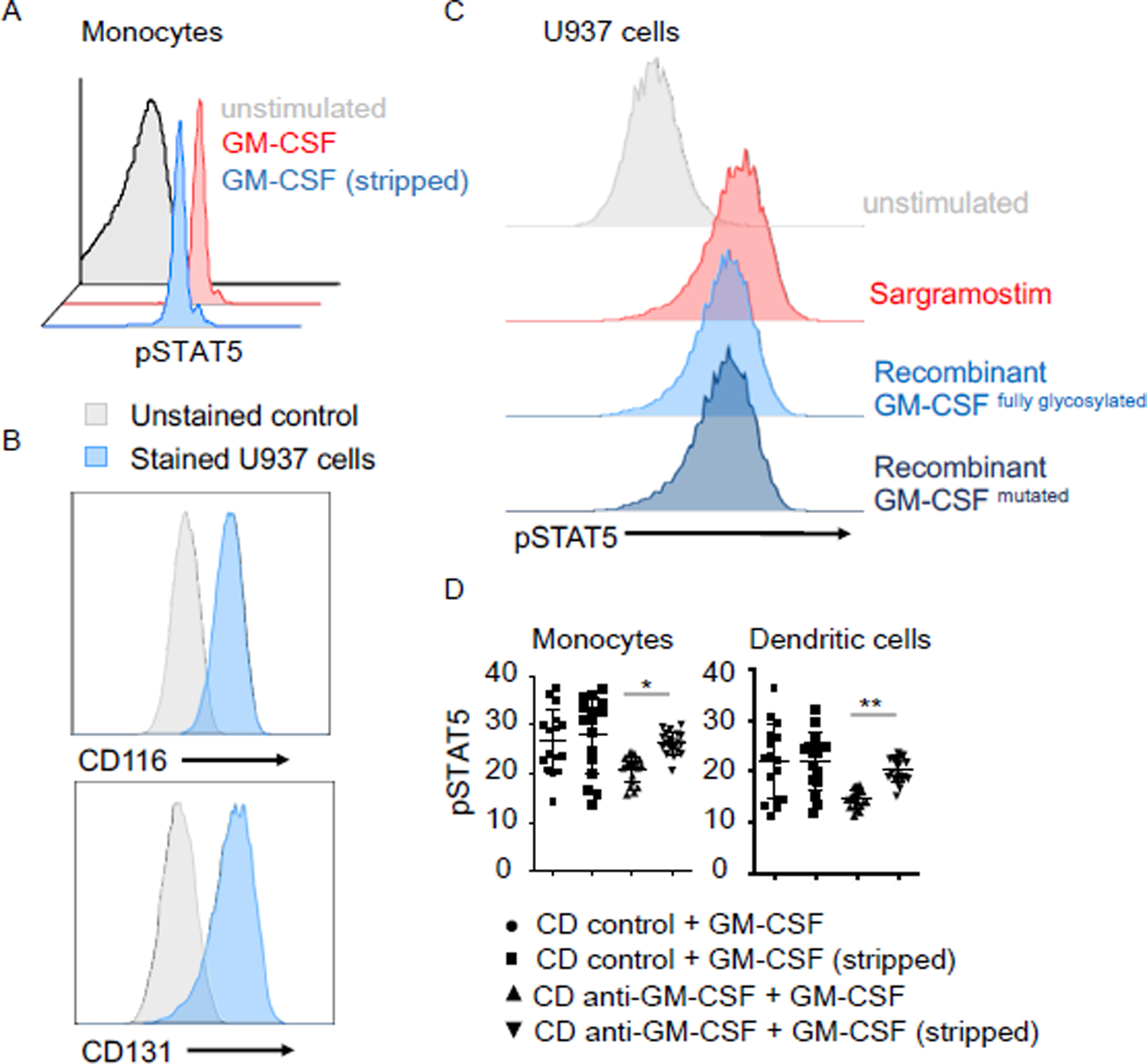
Deglycosylated GM-CSF escapes aGMAb neutralization. **A)** Purified CD14^+^ monocytes were stimulated with rhGM-CSF (sargramostim) or stripped rhGM-CSF for 20min. Cells were analyzed for pSTAT5 levels. **B)** U937 myelomonocytic cells were analyzed for their expression of CD116 and CD131. Histograms show surface stained cells (blue) and unstained controls (grey). **C)** U937 cells were stimulated with rhGM-CSF (sargramostim), purified fully glycosylated GM-CSF or mutated GM-CSF lacking all posttranslational glycosylation sites. Following stimulation, pSTAT5 levels were analyzed. **D)** Plots show quantification of pSTAT5 signal intensity in monocytes and DC either stimulated with GM-CSF or stripped GM-CSF for 20 minutes pre-incubated with serum form the indicated patient groups. One-way analysis of variance (ANOVA) Bonferroni’s multiple comparison test was performed.

**Supplementary Table 1.**
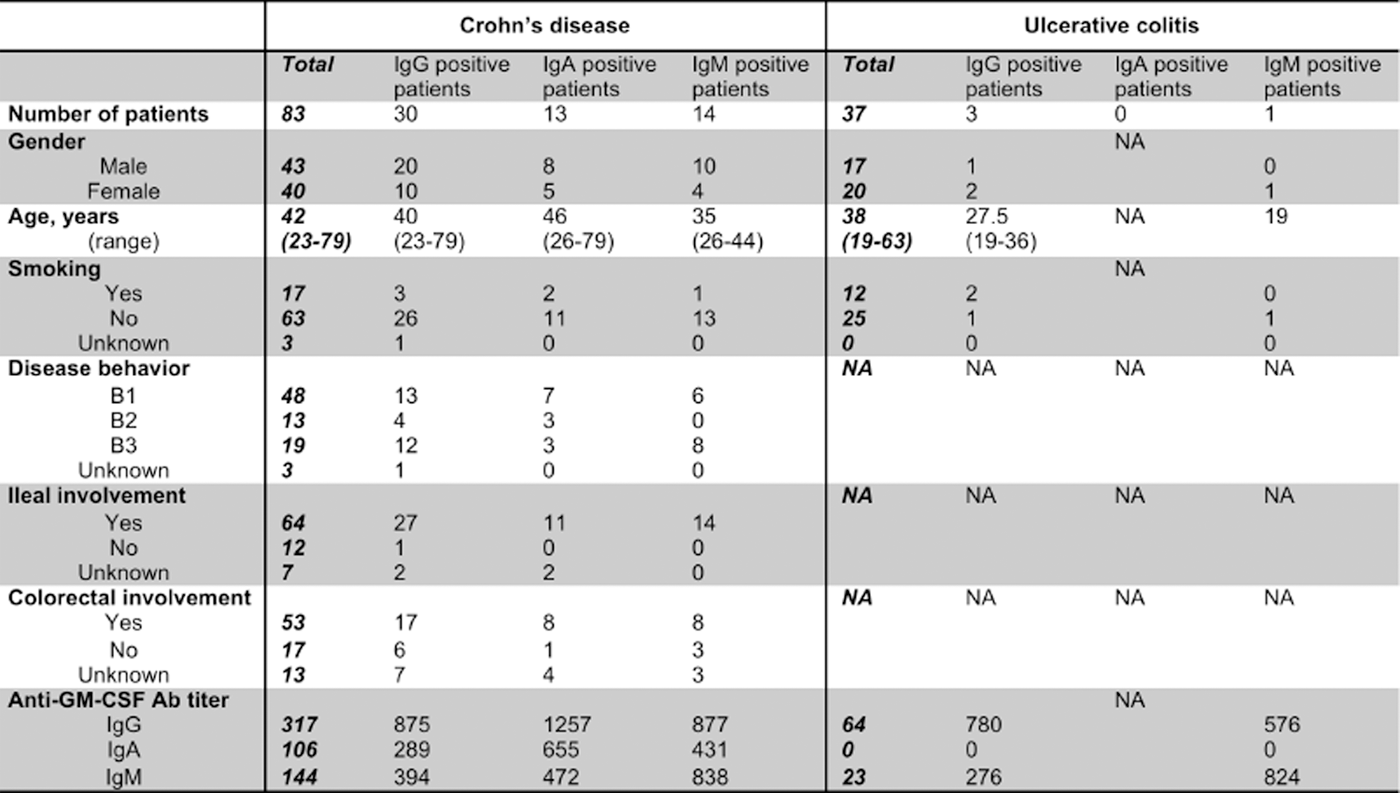
Available patient information for serum samples used in FIGURE.1.

|  | Crohn's disease |  |  |  | Ulcerative colitis |  |  |  |
| --- | --- | --- | --- | --- | --- | --- | --- | --- |
|  | ***Total*** | IgG positive patients | IgA positive patients | IgM positive patients | ***Total*** | IgG positive patients | IgA positive patients | IgM positive patients |
| **Number of patients** | **83** | 30 | 13 | 14 | **37** | 3 | 0 | 1 |
| **Gender** |  |  |  |  |  |  | NA |  |
| Male | **43** | 20 | 8 | 10 | **17** | 1 |  | 0 |
| Female | **40** | 10 | 5 | 4 | **20** | 2 |  | 1 |
| **Age, years** | **42** | 40 | 46 | 35 | **38** | 27.5 | NA | 19 |
| (range) | **(23-79)** | (23-79) | (26-79) | (26-44) | **(19-63)** | (19-36) |  |  |
| **Smoking** |  |  |  |  |  |  | NA |  |
| Yes | **17** | 3 | 2 | 1 | **12** | 2 |  | 0 |
| No | **63** | 26 | 11 | 13 | **25** | 1 |  | 1 |
| Unknown | **3** | 1 | 0 | 0 | **0** | 0 |  | 0 |
| **Disease behavior** |  |  |  |  | **NA** | NA | NA | NA |
| B1 | **48** | 13 | 7 | 6 |  |  |  |  |
| B2 | **13** | 4 | 3 | 0 |  |  |  |  |
| B3 | **19** | 12 | 3 | 8 |  |  |  |  |
| Unknown | **3** | 1 | 0 | 0 |  |  |  |  |
| **Ileal involvement** |  |  |  |  | **NA** | NA | NA | NA |
| Yes | **64** | 27 | 11 | 14 |  |  |  |  |
| No | **12** | 1 | 0 | 0 |  |  |  |  |
| Unknown | **7** | 2 | 2 | 0 |  |  |  |  |
| **Colorectal involvement** |  |  |  |  | **NA** | NA | NA | NA |
| Yes | **53** | 17 | 8 | 8 |  |  |  |  |
| No | **17** | 6 | 1 | 3 |  |  |  |  |
| Unknown | **13** | 7 | 4 | 3 |  |  |  |  |
| **Anti-GM-CSF Ab titer** |  |  |  |  |  |  | NA |  |
| IgG | **317** | 875 | 1257 | 877 | **64** | 780 |  | 576 |
| IgA | **106** | 289 | 655 | 431 | **0** | 0 |  | 0 |
| IgM | **144** | 394 | 472 | 838 | **23** | 276 |  | 824 |

**Supplementary Table 2.** Table shows antibodies and isotope conjugates used in mass-cytometry analysis of peripheral blood mononucleated cells (PBMC) and lamina propria leukocytes (LPL). Scheme adjacent to tables demonstrate the workflow for pSTAT5 staining in these samples.

| PBMC |  |
| --- | --- |
| isotope | antigen |
| Pd102Di | Pd102Di |
| Rh103Di | Viability |
| Pd104Di | Pd104Di |
| Pd105Di | Pd105Di |
| Pd106Di | Pd106Di |
| Pd108Di | Pd108Di |
| Pd110Di | Pd110Di |
| Nd144Di | CD141 |
| Nd148Di | CD16 |
| Nd150Di | pSTAT5 |
| Eu151Di | CD123 |
| Sm152Di | CD66b |
| Eu153Di | CD1c |
| Tb159Di | CD45_159 |
| Gd160Di | CD14 |
| Dy162Di | CD64 |
| Ho165Di | CD45_165 |
| Tm169Di | CD45_169 |
| Er170Di | CD3 |
| Yb172Di | CD11b |
| Yb173Di | CD56 |
| Yb174Di | HLA-DR |
| Lu175Di | CD45_175 |
| Ir191Di | DNA |
| Ir193Di | DNA |

B
| LPL |  |
| --- | --- |
| isotope | antigen |
| Rh103Di | viability |
| In115Di | CD45_115 |
| La139Di | FceR1a |
| Pr141Di | CD45_141 |
| Nd142Di | CD19 |
| Nd144Di | CD141 |
| Nd145Di | CD4 |
| Nd146Di | CD8 |
| Nd148Di | CD16 |
| Sm149Di | CD66 |
| Nd150Di | p-STAT5 |
| Eu151Di | CD123 |
| Eu153Di | CD1c |
| Sm154Di | CD163 |
| Tb159Di | CD11c |
| Gd160Di | CD14 |
| Dy162Di | CD32 |
| Ho165Di | CD116 |
| Er166Di | CD24 |
| Er167Di | CD38 |
| Er168Di | CD206 |
| Tm169Di | CD131 |
| Er170Di | CD3 |
| Yb173Di | CD56 |
| Yb174Di | HLADR |
| Ir191Di | DNA |
| Ir193Di | DNA |

**Supplementary Table 3.** Available patient information for serum samples used in Figure.3, Figure S3 and Figure S4.

|  | Training cohort |  |  | Validation cohort |  |  |
| --- | --- | --- | --- | --- | --- | --- |
|  | CD | UC | HC | CD | UC | HC |
| N subjects | 120 | 100 | 100 | 100 | 100 | 100 |
| N samples | 360 | 300 | 300 | 400 | 400 | 400 |
| Mean age | 30.8 | 29.11 | 29.18 | 31.9 | 32.2 | 34.39 |
| Sex (M) | 79% | 100% | 100% | 78% | 80% | 97% |
| % complication | 28.3% | - | - | 28% | - | - |
| L1 | 27.5 |  |  | 11% | - | - |
| L2 | 15.8 |  |  | 33% | - | - |
| L3 | 39.1 |  |  | 30% | - | - |
| Unknown location | 17.5 |  |  | 26% | - | - |

**Supplementary Table 4.** ELISAs for ASCA-specific antibodies were performed on serum samples collected at three different time points (time point 1 and 2 = prior to disease diagnosis, time point 3 post disease diagnosis). ASCA-specific IgG and IgA were measured.

|  | Healthy Controls | Ulcerative Colitis | Crohn’s Disease |
| --- | --- | --- | --- |
| ASCA-IgG (-2000 days) | Spearman r = 0.40; p < 0.0001 **** | Spearman r = 0.35; p = 0.0003 *** | Spearman r = 0.56; p < 0.0001 **** |
| ASCA-IgG (-500 days) | Spearman r = 0.38; p < 0.0001 **** | Spearman r = 0.31; p = 0.0019 ** | Spearman r = 0.57; p < 0.0001 **** |
| ASCA-IgG (+100 days) | Spearman r = 0.48; p < 0.0001 **** | Spearman r = 0.29; p = 0.0034 ** | Spearman r = 0.70; p < 0.0001 **** |
| ASCA-IgA (-2000 days) | Spearman r = 0.18; n.s. | Spearman r = 0.18; n.s. | Spearman r = 0.17; n.s. |
| ASCA-IgA (-500 days) | Spearman r = 0.15; n.s. | Spearman r = 0.18; n.s. | Spearman r = 0.23; p = 0.0115 * |
| ASCA-IgA (+ 100 days) | Spearman r = N/A; n.s. | Spearman r = 0.17; n.s. | Spearman r = 0.26; p = 0.0049 ** |

## Graphical abstract: Establishing a pre-diseased state through aGMAb in CD

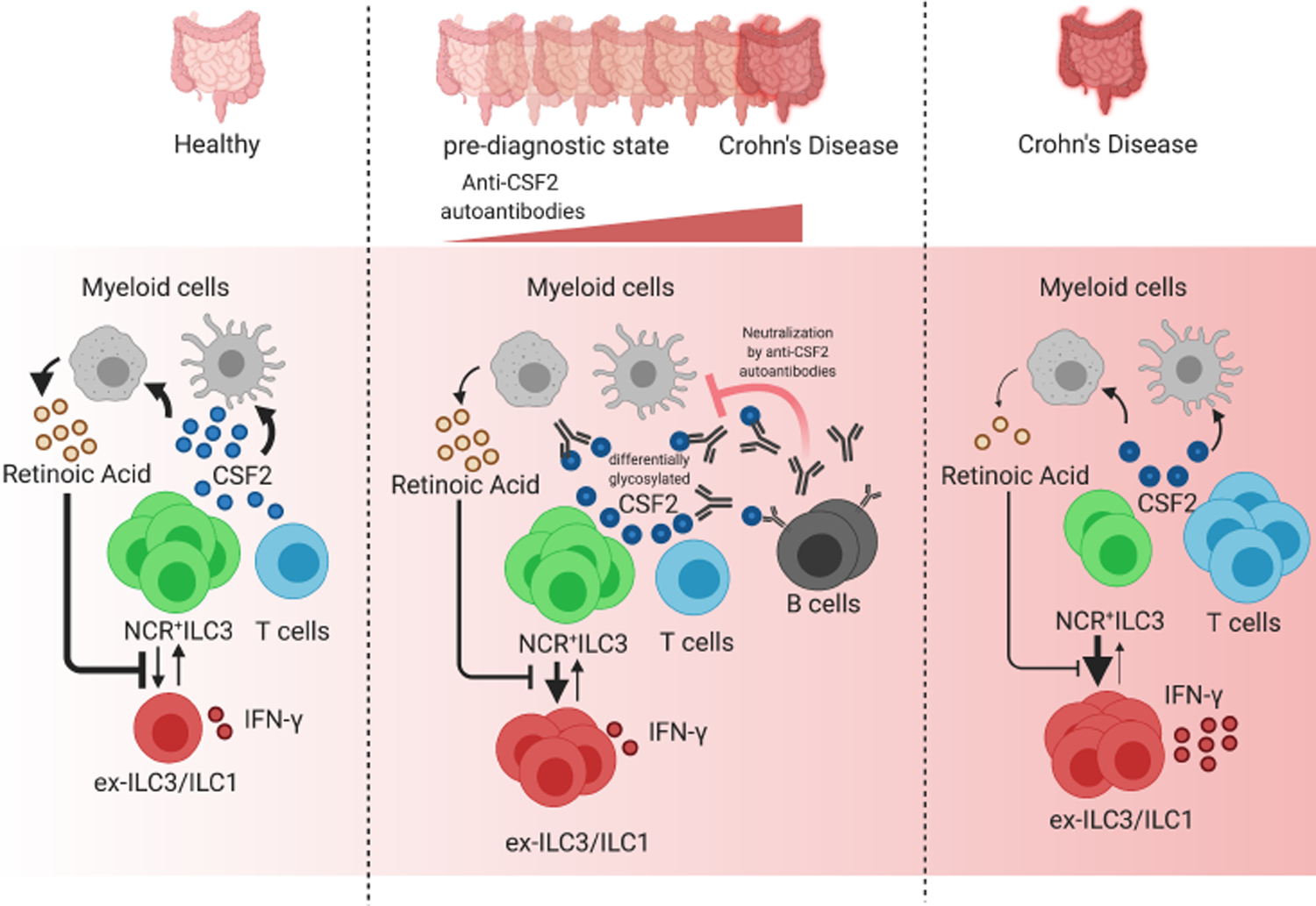
Model of CD development in aGMAb^+^ individuals. Scheme shows cellular crosstalk in healthy intestinal tissue (left). NCR^+^ILC3 and T cells produce GM-CSF that engages the GM-CSFR on myeloid cells to trigger the production of RA. Retinoic acid in turn stabilizes NCR^+^ILC3 and prevents excessive differentiation of NCR^+^ILC3 into IFN-γ producing ILC1. The middle scheme displays the scenario in the presence of aGMAb. GM-CSF produced by T cells and NCR^+^ILC3 is aberrantly glycosylated and may drive the activation of glycosylation-specific B cells. B cell-derived aGMAb neutralize GM-CSF and reduce GM-CSFR signaling, leading to a decreased production of retinoic acid. Consequently, NCR^+^ILC3s differentiation into IFNγ producing ILC1. The right scheme resembles the immune landscape in full-blown CD. The inflamed CD mucosa is characterized by a decrease in NCR^+^ILC3 numbers, lower levels of GM-CSF produced by NCR^+^ILC3, reduced levels of RA from myeloid cells and excessive differentiation into IFNγ producing ILC1 and T cells. The presence of aGMAb may interrupt the NCR^+^ILC3 – myeloid cell circuit, destabilizing homeostasis towards the scenario of full-blown CD.

